# An 81 base-pair deletion in SARS-CoV-2 ORF7a identified from sentinel surveillance in Arizona (Jan-Mar 2020)

**DOI:** 10.1101/2020.04.17.20069641

**Authors:** LaRinda A. Holland, Emily A. Kaelin, Rabia Maqsood, Bereket Estifanos, Lily I. Wu, Arvind Varsani, Rolf U. Halden, Brenda G. Hogue, Matthew Scotch, Efrem S. Lim

**Author notes:** These authors equally contributed to this work (LaRinda A. Holland, Emily A. Kaelin).

## Abstract

On January 26 2020, the first Coronavirus Disease 2019 (COVID-19) case was reported in Arizona of an individual with travel history (3^rd^ case in the US) (1). Here, we report on early SARS-CoV-2 sentinel surveillance in Tempe, Arizona (USA). Genomic characterization identified an isolate encoding a 27 amino acid in-frame deletion in accessory protein ORF7a, the ortholog of SARS-CoV immune antagonist ORF7a/X4.

---

In anticipation of COVID-19 spreading in the state of Arizona, we initiated a surveillance effort for local emergence of SARS-CoV-2 starting January 24, 2020. We leveraged an ongoing influenza surveillance project at Arizona State University (ASU) Health Services in Tempe, Arizona. Individuals presenting with respiratory symptoms (ILI) were tested for influenza A and B virus (Alere BinaxNOW). Subsequently, we tested influenza-negative nasopharyngeal (NP) swabs for SARS-CoV-2. We extracted total nucleic acid using the bioMérieux eMAG automated platform and performed real-time RT-PCR (qRT-PCR) assays specific for SARS-CoV-2 N and E genes (2, 3). Out of 382 NP swabs collected from January 24, 2020 to March 25, 2020, we detected SARS-CoV-2 in 5 swabs in the week of March 16 to 19 (**Figure 1**). This corresponds to prevalence of 1.31%. Given the estimated 1 – 14-day incubation period for COVID-19, it is possible that the spike in cases might be related to university spring-break holiday travel (March 8 – 15) as previously seen in other outbreaks (4, 5).

**Figure 1:**
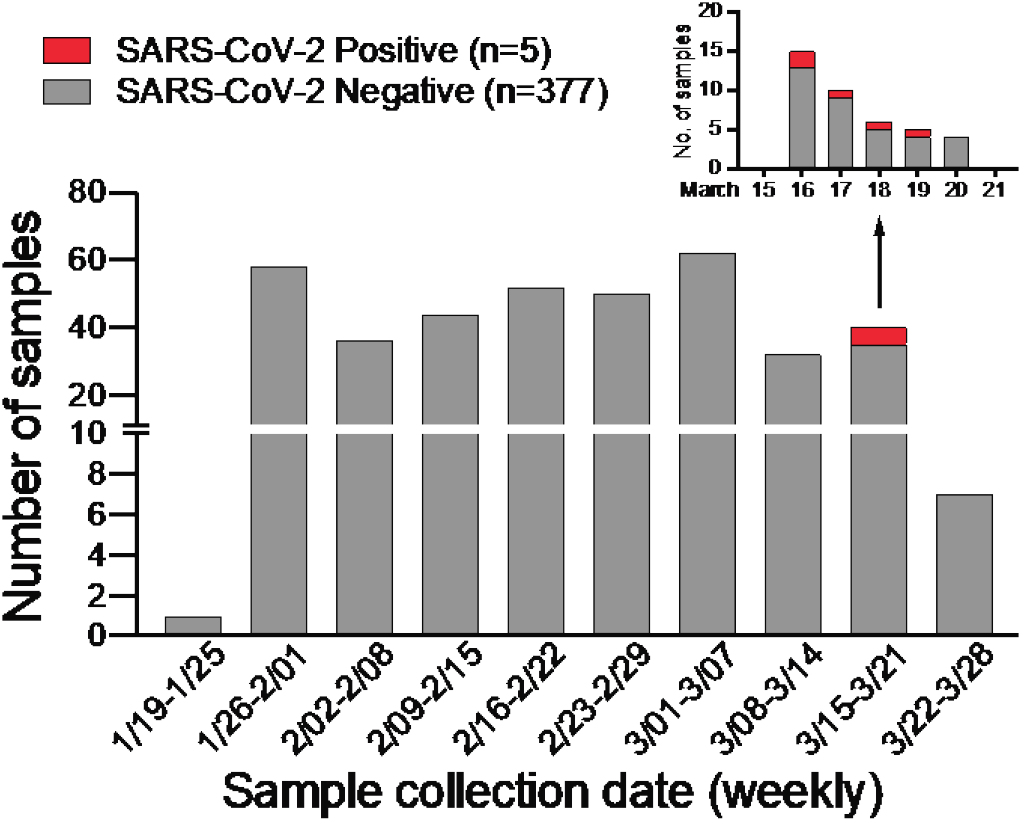
SARS-CoV-2 surveillance in Tempe, AZ from January to March 2020. Weekly distribution of NP specimens collected by ASU Health Services tested for SARS-CoV-2 by qRT-PCR assays. Inset shows SARS-CoV-2 positive NP specimens collected from the week of March 15 – 21, 2020.

To understand the evolutionary relationships and characterize the SARS-CoV-2 genomes, we performed next-generation sequencing (Illumina NextSeq, 2×76) directly on specimen RNA, thereby avoiding cell culture passage and potentially associated mutations. This generated an NGS dataset of 20.7 to 22.7 million paired-end reads per sample. We mapped quality-filtered reads to a reference SARS-CoV-2 genome (MN908947) using BBMap (version 39.64) to generate three full-length genomes: AZ-ASU2922 (376x coverage), AZ-ASU2923 (50x) and AZ-ASU2936 (879x) (Geneious prime version 2020.0.5). We aligned a total of 222 SARS-CoV-2 genome sequences comprising at least 5 representative sequences from phylogenetic lineages defined by Rambaut *et al*. (6), ranging from January 5 to March 31, 2020 from 25 different countries. We performed phylogenetic reconstruction with BEAST (version 1.10.4, strict molecular clock, HKY + Γ nucleotide substitution, exponential growth for coalescent model) (7-10). The ASU sequences were phylogenetically distinct indicating that they were independent transmissions (**Figure 2A**).

**Figure 2:**
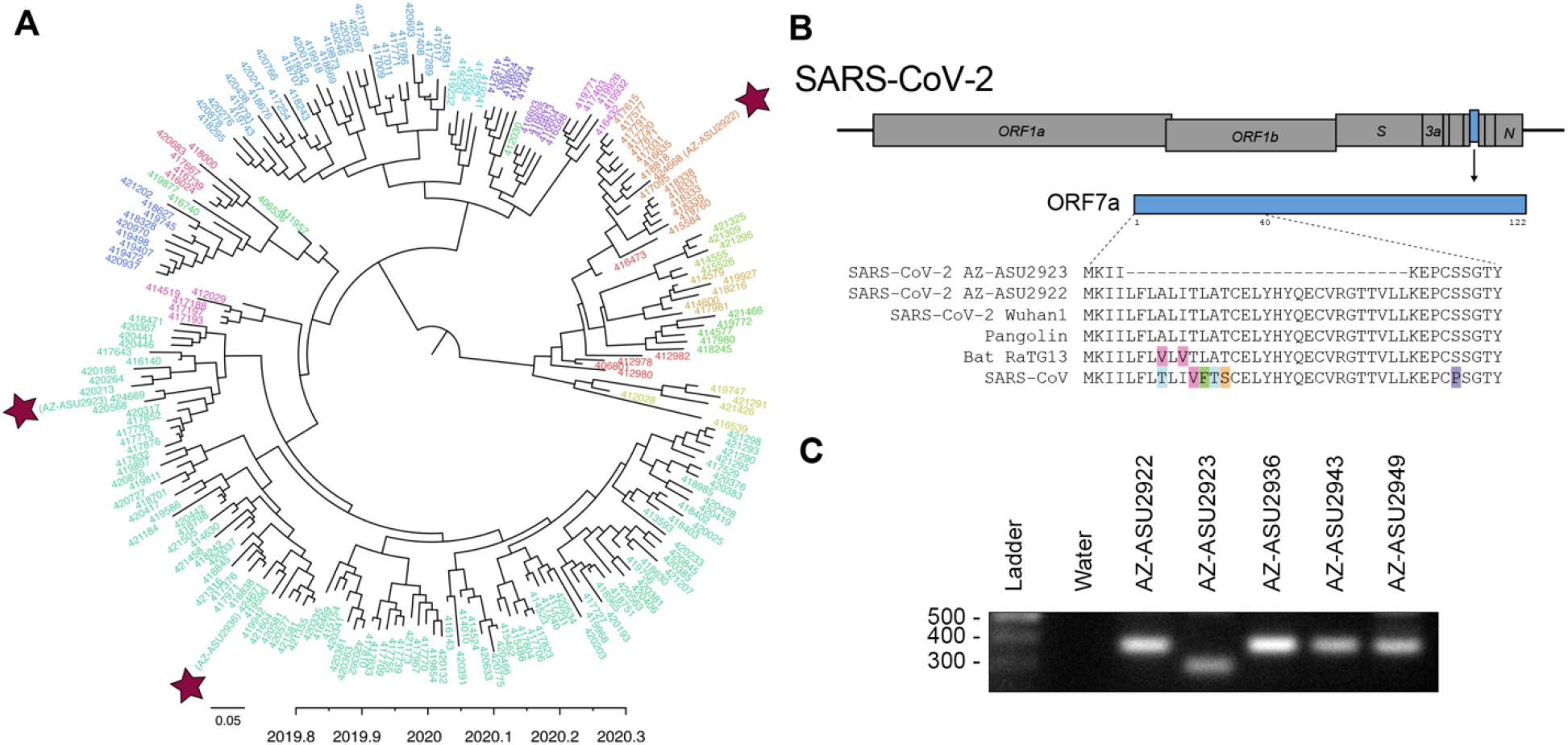
Evolutionary and genomic characterization of SARS-CoV-2 genomes. (**A**) Bayesian maximum clade credibility (MCC) polar phylogeny of 222 full-length SARS-CoV-2 genomes. The 3 new genomes reported in this study are indicated by red stars. Sequenced were aligned in Geneious prime (version 2020.0.3) using the MAFFT v7.450 plugin, and trimmed the 5’ and 3’ UTR (< 300 nts each). We initiated two independent runs of 500M sampling every 50K steps and used Tracer v1.7.1 (16) to check for convergence and that all ESS values for our statistics were > 200, LogCombiner (7) to combine the models with a 10% burn-in and TreeAnnotator (7) to produce a MCC tree. We used FigTree v1.4.4 (17) to edit the tree and color the tips based on lineages (6), and pangolin (18) to identify the lineages of our 3 new sequences based on the established nomenclature (6). The nomenclature consists of two main lineages, A and B, and includes “sub-lineages” (A.1, B.2. *etc*.) up to four levels deep (e.g. A.1.1, B.2.1) (6). For visualization purposes, we grouped all viruses that were not directly assigned to “A” or “B” into their first sub-lineage level and colored tip labels by lineage. B.1 lineage: AZ-ASU2923 and AZ-ASU2936; A.1 lineage AZ-ASU2922. (**B**) ORF7a amino acid alignment of SARS-CoV-2 and related genomes. GenBank and GISAID accession numbers: SARS-CoV-2 AZ-ASU2922 (MT339039, EPI_ISL_424668), SARS-CoV-2 AZ-ASU2923 (MT339040, EPI_ISL_424669), SARS-CoV-2 AZ-ASU2936 (MT339041, EPI_ISL_424671), SARS-CoV-2 Wuhan1 (MN908947.3), Pangolin (EPI_ISL_410721), Bat-RaTG13 (MN996532.1), SARS-CoV (AY278741.1). The 81-bp (27 amino acid) deletion observed in SARS-CoV-2 AZ_ASU2923 ORF7a was not present in the 6,290 SARS-CoV-2 sequences available from GISAID as of April 12, 2020. (**C**) We performed molecular validation by RT-PCR on specimen total nucleic acid extracts with primers flanking the ORF7a N-terminus region. The expected size of amplicons with intact ORF7a region is 377bp, the expected size of an amplicon with the NGS-identified 81bp deletion is 296bp. Primers: SARS2-27144F 5’-ACAGACCATTCCAGTAGCAGTG-3’, SARS2-27520r 5’-TGCCCTCGTATGTTCCAGAAG-3’.

Similar to SARS-CoV, the SARS-CoV-2 genome encodes multiple open reading frames in the 3’ region. We found that the SARS-CoV-2 AZ-ASU2923 genome has an 81 base-pair deletion in the ORF7a gene resulting in a 27 amino-acid in-frame deletion (**Figure 2B**). The SARS-CoV ORF7a ortholog is a viral antagonist of host restriction factor BST-2/Tetherin and induces apoptosis (11-14). Based on the SARS-CoV ORF7a structure (15), the 27-aa deletion in SARS-CoV-2 ORF7a maps to the putative signal peptide (partial) and first two beta strands. To validate the deletion, we performed RT-PCR using primers spanning the region and verified by Sanger sequencing the amplicons (**Figure 2C and Supplementary Figure 1**).

Collectively, although global NGS efforts indicate that SARS-CoV-2 genomes are relatively stable, dynamic mutations can be selected in symptomatic individuals.

## Data Availability

Sequence data has been deposited to NCBI GenBank (MT339039, MT339040 and MT339041) and GISAID (EPI_ISL_424668, EPI_ISL_424669 and EPI_ISL_424671).

## Acknowledgements

We thank the nurses and staff at the ASU Health Services, Arizona Department of Health Services for a SARS-CoV-2 positive sample (AZ_4811) for qRT-PCR assay validation experiments, Nicholas Mellor and the ASU Genomics Facility for technical assistance, the authors, originating and submitting laboratories of the sequences from GISAID’s EpiCoV™ Database. A complete acknowledgements table is available at https://www.dropbox.com/s/aiybuatgxjunuga/GISAID_CoV2020_Acknowledgements.xlsx?dl=0. This work was supported by NSF STC Award 1231306 (B.G.H), NIH grants R01 LM013129 (R.U.H., A.V., M.S.), R00 DK107923 (E.S.L.), J.M. Kaplan Foundation’s One Water One Health (Arizona State University Foundation project 30009070) and ASU Core Facilities Seed Funding.

## Data availability

Sequence data has been deposited to NCBI GenBank and GISAID: SARS-CoV-2 AZ-ASU2922 (MT339039, EPI_ISL_424668), SARS-CoV-2 AZ-ASU2923 (MT339040, EPI_ISL_424669) and SARS-CoV-2 AZ-ASU2936 (MT339041, EPI_ISL_424671).

## Figure legends

**Supplementary Figure 1.**
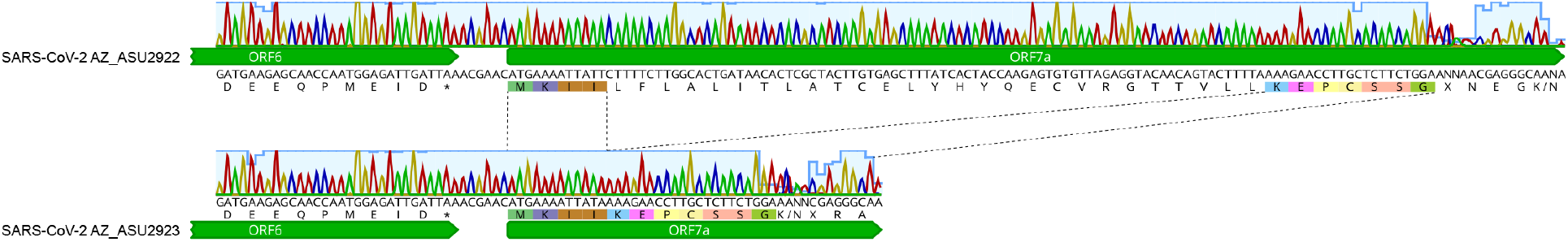
Sanger sequencing chromatograms verification of 81-bp deletion in SARS-CoV-2 AZ-ASU2923 in the ORF7a region.

## Notes

### Competing Interest Statement

The authors have declared no competing interest.

### Funding Statement

This work was supported by NSF STC Award 1231306 (B.G.H), NIH grants R01 LM013129 (R.U.H., A.V., M.S.), R00 DK107923 (E.S.L.) and ASU Core Facilities Seed Funding.

## References

1. Patel A, Jernigan DB. 2020. Initial Public Health Response and Interim Clinical Guidance for the 2019 Novel Coronavirus Outbreak - United States, December 31, 2019-February 4, 2020. MMWR Morb Mortal Wkly Rep 69:140–146.

2. CDC. 2020. 2019-Novel Coronavirus (2019-nCoV) Real-time rRT-PCR Panel Primers and Probes. https://www.cdc.gov/coronavirus/2019-ncov/downloads/rt-pcr-panel-primer-probes.pdf Accessed 3/4/2020.

3. Corman VM, Landt O, Kaiser M, Molenkamp R, Meijer A, Chu DKW, Bleicker T, Brunink S, Schneider J, Schmidt ML, Mulders D, Haagmans BL, van der Veer B, van den Brink S, Wijsman L, Goderski G, Romette JL, Ellis J, Zambon M, Peiris M, Goossens H, Reusken C, Koopmans MPG, Drosten C. 2020. Detection of 2019 novel coronavirus (2019-nCoV) by real-time RT-PCR. Euro Surveill 25.

4. Lauer SA, Grantz KH, Bi Q, Jones FK, Zheng Q, Meredith HR, Azman AS, Reich NG, Lessler J. 2020. The Incubation Period of Coronavirus Disease 2019 (COVID-19) From Publicly Reported Confirmed Cases: Estimation and Application. Ann Intern Med doi:10.7326/m20-0504.

5. Polgreen PM, Bohnett LC, Yang M, Pentella MA, Cavanaugh JE. 2010. A spatial analysis of the spread of mumps: the importance of college students and their spring-break-associated travel. Epidemiol Infect 138:434–41.

6. Rambaut A, Holmes EC, Pybus OG. 2020. A dynamic nomenclature for SARS-CoV-2 to assist genomic epidemiology. http://virological.org/t/a-dynamic-nomenclature-for-sars-cov-2-to-assist-genomic-epidemiology/458.

7. Suchard MA, Lemey P, Baele G, Ayres DL, Drummond AJ, Rambaut A. 2018. Bayesian phylogenetic and phylodynamic data integration using BEAST 1.10. Virus Evol 4:vey016.

8. Hasegawa M, Kishino H, Yano T-a. 1985. Dating of the human-ape splitting by a molecular clock of mitochondrial DNA. Journal of Molecular Evolution 22:160–174.

9. Yang Z. 1994. Maximum likelihood phylogenetic estimation from DNA sequences with variable rates over sites: Approximate methods. Journal of Molecular Evolution 39:306–314.

10. Pybus OG, Rambaut A. 2002. GENIE: estimating demographic history from molecular phylogenies. Bioinformatics 18:1404–1405.

11. Taylor JK, Coleman CM, Postel S, Sisk JM, Bernbaum JG, Venkataraman T, Sundberg EJ, Frieman MB. 2015. Severe Acute Respiratory Syndrome Coronavirus ORF7a Inhibits Bone Marrow Stromal Antigen 2 Virion Tethering through a Novel Mechanism of Glycosylation Interference. J Virol 89:11820–33.

12. Yuan X, Wu J, Shan Y, Yao Z, Dong B, Chen B, Zhao Z, Wang S, Chen J, Cong Y. 2006. SARS coronavirus 7a protein blocks cell cycle progression at G0/G1 phase via the cyclin D3/pRb pathway. Virology 346:74–85.

13. Tan YJ, Fielding BC, Goh PY, Shen S, Tan TH, Lim SG, Hong W. 2004. Overexpression of 7a, a protein specifically encoded by the severe acute respiratory syndrome coronavirus, induces apoptosis via a caspase-dependent pathway. J Virol 78:14043–7.

14. Schaecher SR, Touchette E, Schriewer J, Buller RM, Pekosz A. 2007. Severe acute respiratory syndrome coronavirus gene 7 products contribute to virus-induced apoptosis. J Virol 81:11054–68.

15. Nelson CA, Pekosz A, Lee CA, Diamond MS, Fremont DH. 2005. Structure and intracellular targeting of the SARS-coronavirus Orf7a accessory protein. Structure 13:75–85.

16. Rambaut A, Drummond AJ, Xie D, Baele G, Suchard MA. 2018. Posterior Summarization in Bayesian Phylogenetics Using Tracer 1.7. Syst Biol 67:901–904.

17. Rambaut A. 2018. FigTree v1.4.4, https://github.com/rambaut/figtree.

18. O’Toole A, McCrone J. 2020. pangolin, https://github.com/hCoV-2019/pangolin.

